# Comparative impacts and cost-effectiveness of tuberculosis active case-finding strategies in prisons in Brazil, Colombia, and Peru: a mathematical modeling study

**DOI:** 10.1101/2025.09.25.25336625

**Authors:** Yiran E Liu, José Victor Bortolotto Bampi, Ronan F Arthur, Argita D Salindri, Caroline Busatto, Pedro Avedillo Jiménez, Daniele Maria Pelissari, Fernanda Dockhorn Costa Johansen, Robert Arana-Narvaez, Alvaro Fernando Moreno Roca, Wilfredo Santos Solís Tupes, Esther Mori Jiu, Christian Alfredo Moreno Roca, Erika Albertina Abregú Contreras, Valentina Antonieta Alarcón Guizado, Julián Trujillo Trujillo, Belkys Marcelino, Mónica Alonso Gonzalez, Mayra Cecilia Córdova Ayllon, Ted Cohen, Moises A Huaman, Jeremy D Goldhaber-Fiebert, Julio Croda, Jason R Andrews

## Abstract

**Background:** Incarceration is a leading driver of tuberculosis in Latin America. Active case-finding in prisons may reduce population-wide tuberculosis burden, but optimal strategies and cost-effectiveness remain uncertain.

**Methods and findings:** Using dynamic transmission models calibrated to Brazil, Colombia, and Peru, we simulated annual or biannual (twice-yearly) prison-wide screening, alone or combined with entry and exit screening from 2026-2035. We evaluated four algorithms: 1) symptom screening, 2) chest X-ray with computer-aided detection (CXR-CAD), 3) symptoms and CXR-CAD (follow-up testing if either is positive) and 4) GeneXpert Ultra with pooled sputum. Individuals screening positive then received individual Xpert. We projected impacts on within-prison and population-level tuberculosis incidence in 2035, along with discounted costs (2023 USD) and disability-adjusted life years (DALYs). Model projections showed that combined entry, exit, and biannual screening with CXR-CAD was highly impactful and cost-effective across countries, reducing tuberculosis incidence by 62-87% in prisons and 18-28% population-wide. Compared to only biannual CXR-CAD (the next best strategy), the incremental cost per DALY averted of adding entry and exit screening was $2984 (Brazil), $2925 (Colombia), and $645 (Peru). Adding symptom screening to CXR-CAD marginally increased benefit and was only cost-effective in Peru’s higher-incidence prisons. Biannual screening alone remained cost-effective at prison incidence levels well below national averages. In settings without CXR-CAD, pooled Xpert was an impactful, cost-effective alternative.

**Conclusions:** These modeling results support immediate national-level adoption of prison-wide tuberculosis screening twice-yearly and at entry and exit. Screening should begin with available methods while building capacity for CXR-CAD, the most cost-effective algorithm.

**AUTHOR SUMMARY:** *Why was this study done?:*

- In Latin America, rising incarceration has fueled the tuberculosis epidemic, with extremely high infection rates among people deprived of liberty. These effects extend beyond prison walls, driving tuberculosis spread in outside communities.
- Interventions targeted to prisons may have an outsized impact on reducing tuberculosis in the broader population.
- The World Health Organization strongly recommends systematic screening for tuberculosis in prisons, but there is little evidence on how often to screen, which methods to use, and whether these approaches are cost-effective across different country contexts.

*What did the researchers do and find?:*

- We developed mathematical models using data from Brazil, Colombia, and Peru to simulate different prison-based tuberculosis screening strategies and project their health impacts and costs.
- We compared prison-wide screening once or twice a year, screening at prison entry or exit, and combinations of these approaches. We also compared different screening methods using symptoms, chest X-ray with computer-aided detection (CXR-CAD), or pooled molecular testing (GeneXpert Ultra).
- The models projected that combining entry, exit, and twice-yearly prison-wide screening with CXR-CAD would be highly impactful and cost-effective in all three countries. This strategy could substantially reduce tuberculosis in prisons and in the general population.
- Twice-yearly prison-wide screening remained cost-effective even in prisons with much lower tuberculosis rates than national averages.
- CXR-CAD was the optimal screening method, but pooled molecular testing was also impactful and cost-effective where CXR-CAD was not available.

*Implications of all the available evidence:*

- Systematic screening in prisons, twice-yearly and at entry and exit, is projected to be highly impactful and cost-effective across different settings in Latin America.
- These findings support urgent adoption of intensive prison-based tuberculosis screening throughout the region, starting with the best available diagnostic tools while investing in CXR-CAD.

## INTRODUCTION

Tuberculosis persists as a leading global health threat, with an estimated 10.8 million people falling ill and 1.25 million people dying from the disease in 2023^1^. People deprived of liberty (PDL) experience substantially elevated risk of tuberculosis due to prison conditions like overcrowding, poor ventilation, malnutrition, and limited access to medical care^2^. Timely diagnosis and treatment are an essential component of tuberculosis control in prisons, but studies estimate that only about half of PDL with tuberculosis globally are diagnosed^3^.

In Latin America, tuberculosis incidence and deaths have increased since 2015, and the worsening tuberculosis epidemic is increasingly driven by prisons^2^. Our recent modeling study estimated that, following decades of rising incarceration rates, 27% of new cases in the region were attributable to incarceration^4^. Interventions to address tuberculosis in prisons may have an outsized impact on the population-wide tuberculosis epidemic in Latin America.

Tuberculosis active case-finding, or systematic screening, may improve early detection and reduce transmission in prisons^5^. Since 2021, the World Health Organization (WHO) has strongly recommended systematic screening in prisons, at minimum annually and upon entry and exit^6^. However, the guidelines cite a “very low certainty of evidence” and highlight the need for more research on the effectiveness and cost-effectiveness of different strategies. Recent empirical studies in prisons have demonstrated high yield from strategies using chest x-ray with computer-automated detection (CXR-CAD) or molecular rapid testing (i.e. GeneXpert) with sputum pooling^7–10^. However, their broader impact and cost-effectiveness remain uncertain, especially across settings with varying tuberculosis epidemiology and carceral characteristics^6^. Moreover, the WHO has described biannual (twice-yearly) mass screening in prisons as “ideal,” but lacking economic evidence, cited resource constraints as potential barriers to sustained implementation^11^. Existing modeling studies in the region offer limited guidance: of two prior studies, both were conducted in Brazil only and used subnational data, and neither assessed cost-effectiveness nor modeled screening more than once per year^12,13^.

In this study, we utilize a compartmental dynamic transmission model to project the population-level impact and cost-effectiveness of tuberculosis active case-finding interventions in prisons in Brazil, Colombia, and Peru. We evaluate strategies for screening across different timepoints (i.e., entry, exit, periodic), frequencies (annual or biannual), and algorithms (symptom screening, CXR-CAD, combined symptoms and CXR-CAD, or GeneXpert Ultra with sputum pooling). All algorithms include follow-up testing with individual GeneXpert Ultra. We also perform sensitivity analyses to identify preferred strategies in settings with lower prison incidence or without access to CXR-CAD technology.

## METHODS

### Transmission model and key assumptions

We extended a published compartmental transmission model simulating tuberculosis and incarceration dynamics^4^. The model includes a simple representation of tuberculosis natural history for the population 15 years of age and older. Upon infection, **S**usceptible individuals transition to **E**arly latent infection, after which they activate to **I**nfectious disease or transition to **L**ate latent infection, with lower risk of active disease. Individuals in **I** transition to **R**ecovered through self-cure or diagnosis and successful treatment. Individuals in **L** and **R** can be re-infected to **E**, and individuals in **R** can relapse to **I**.

These natural history compartments are reproduced across four population strata, which individuals traverse upon incarceration, release, and re-incarceration: prison (**p)**, recent history of incarceration (**r)**, distant history of incarceration (**d)**, and never incarcerated (**n**). The model is not stratified by age, gender/sex, or HIV. Multi-drug resistant (MDR) tuberculosis was not modeled explicitly but was accounted for by adjusting treatment outcomes and costs based on relative proportions of drug-sensitive and drug-resistant tuberculosis.

The model was separately calibrated for Brazil, Colombia, and Peru to country-specific data from 1990-2023. Data targets included prevalence of incarceration, prison admission rates, recidivism, population-wide tuberculosis notifications and incidence, and within-prison tuberculosis notifications and incidence^4^. Calibration targets for prison incidence were based on a previous study using Bayesian meta-regression modeling to estimate within-prison case-detection ratios (**Appendix**)^3^. Model fit to data is shown in **Figures S1-S2**. From 2023 on, we assume stable prison entry and release rates, with incarceration rates eventually leveling off. The elevated risk of tuberculosis in prisons is operationalized through higher effective contact rates, higher disease progression rates, and lower diagnosis rates in prisons than in the community^4^.

Individuals with recent incarceration history are assumed to have similar contact rates as never incarcerated individuals, but higher disease progression rates and lower diagnosis rates^4^. The transition from recent to distant incarceration history occurs on average at seven years following release, after which rates of contact, progression, and diagnosis match those of never incarcerated individuals^12^.

### Interventions and scenarios

In the base-case scenario, we assume only passive diagnosis in prisons. Under passive diagnosis, people with presumptive TB undergo clinical evaluation, with a proportion receiving bacteriologic testing using smear microscopy and/or GeneXpert Ultra. A proportion of confirmed cases additionally receive culture and drug susceptibility testing (DST). Coverage of each testing method is informed by country-specific programmatic data **(Table S1)**.

We simulated prison-based active case-finding interventions, in addition to passive diagnosis, implemented from the start of 2026 through the end of 2035. We modeled systematic screening at prison entry or exit, prison-wide screening conducted annually or biannually (twice yearly), or a combination of entry, exit, and annual or biannual screening. We assume entry screening applies to individuals entering prison from the community as well as individuals transferred between prison facilities. Screening of transferred individuals was modeled by adding a continual rate of screening to the prison stratum; movements between prisons were not explicitly modeled. Exit screening only applies to individuals exiting prisons to the community. Parameters governing screening coverage, duration, linkage to care post-release, treatment completion in prison and post-release, and costs are detailed in **Table S1**. Individuals in **E** who test false positive and complete active treatment transition to **L**. Other false positives do not undergo any model-based transitions.

### Screening algorithms and assumptions

For each intervention, we compared four screening algorithms: 1) symptom-based, 2) CXR-CAD, 3) combined symptoms and CXR-CAD, and 4) GeneXpert Ultra MTB/RIF with pooled sputum. We used empirical accuracy estimates from studies of active case-finding in prisons when available. Symptom screening was based on cough of any duration, the symptom for which the most data from prisons are available. For combined symptoms and CXR-CAD, individuals with either symptoms or an abnormal X-ray (or both) are considered screen-positive. For sputum pooling, samples from eight individuals are combined and tested using a single Xpert cartridge; we assume that pooling reduces Xpert sensitivity but does not affect specificity^9^. If the pooled test is positive, each individual sample in the pool is tested separately to identify screen-positive individuals. Individuals who screen positive receive follow-up testing with individual GeneXpert Ultra on a new sputum sample and clinical evaluation. Accuracy and costs of screening methods and follow-up testing are shown in **Table 1**.

**Table 1.** Accuracy and costs of screening interventions. Values listed are the mode (range) from a triangle distribution or the median (range) when values are composites of individual parameters. All costs are in 2023 US dollars (USD) and include consumable and personnel costs. Listed costs of CXR-CAD alone and pooled Xpert also include costs of administering a symptom interview. Follow-up testing includes clinical evaluation and Xpert Ultra. For Xpert pooling, sensitivity is derived from the sensitivity of Xpert Ultra in a screening context, multiplied by the relative sensitivity of using pooled samples. For costs of pooled Xpert, cartridge savings vary based on TB prevalence; listed costs are the median (range) of costs over the ten-year intervention horizon for annual screening. Additional intervention and cost parameters, including costs of individual components, are provided in Table S1. CXR-CAD, chest x-ray with computer-aided detection.

|  | Sensitivity (%) | Specificity (%) | Per-unit cost (2023 USD) |  |  | Source |
| --- | --- | --- | --- | --- | --- | --- |
|  |  |  | Brazil | Colombia | Peru |  |
| Symptom screen | 51 (30-70) | 70 (60-80) | 2.27 (2.00-3.00) | 1.58 (1.39-2.08) | 1.76 (1.50-2.50) | 6,7,20,29,30 |
| CXR-CAD | 85 (75-88.5) | 81 (70-93) | 10.48 (8.28-13.60) | 9.10 (7.40-11.48) | 9.34 (7.24-12.19) | 6,7,10,20 |
| Symptoms & CXR-CAD | 89 (84-93) | 52 (41-63) | 10.48 (8.28-13.60) | 9.10 (7.40-11.48) | 9.34 (7.24-12.19) | 10,31 |
| Pooled Xpert | 66.1 (55.3-78.2) | 98.8 (97-99.5) | 19.38 (13.23-31.12) | 14.01 (9.46-21.61) | 15.62 (10.47-24.49) | 32, 7,33, 34 |
| Follow-up testing | 90.9 (86.2-94.7) | 98 (96-99.5) | 26.43 (19.90-38.38) | 21.93 (16.81-30.86) | 23.50 (17.57-33.06) | 34 |

All individuals diagnosed through screening (including false positives) receive culture, DST, and treatment. Individuals diagnosed through exit screening initiate treatment only if they are linked to care. In any intervention, individuals who initiate but do not complete treatment are assumed to incur half the treatment costs.

### Costs and outcomes

We projected the percent reduction in prison and population tuberculosis incidence in 2035, the last year of the intervention period, compared to the base-case scenario. Across all population strata, we projected disability-adjusted life years (DALYs) due to tuberculosis and post-tuberculosis sequelae^14^ and estimated costs from the health system perspective, including costs of screening, follow-up testing, diagnosis, and treatment for cases detected through passive and active case-finding. We used a ten-year analytic horizon, accounting for lifetime DALY streams. Costs are reported in 2023 US dollars (USD), adjusted for inflation using the World Bank GDP deflator^15^. Costs and DALYS were standardized per 100,000 population and discounted at 3% annually^16^.

Cost parameters were sourced from costing studies conducted in the selected countries (**Appendix**). Costs in the base-case scenario include costs of passive diagnosis and treatment. Intervention costs include screening and treatment. Costs for each screening algorithm include costs of equipment, consumables, and labor for screening and follow-up testing. Costs of treatment include costs of drugs, hospitalization, directly observed treatment (DOT), and follow-up evaluation and testing for the average patient. Treatment costs were assumed to be the same for patients diagnosed through passive and active case-finding. We report costs relative to the base-case scenario as the net of intervention costs and cost savings from averted cases.

For strategies on the cost-effectiveness efficiency frontier, we calculated incremental cost-effectiveness ratios (ICERs), defined as the additional cost for each additional DALY averted compared to the next-best non-dominated strategy^16^. Strategies were considered cost-effective if their ICERs were within country-specific cost-effectiveness thresholds: $8602 (Brazil), $5618 (Colombia), and $5731 (Peru)^17–19^ (**Appendix**).

### Uncertainty and sensitivity analyses

We sampled 1000 sets of intervention parameters and cost inputs from uncertainty distributions using Latin Hypercube sampling. These sets were randomly paired with 1000 sets of fitted model parameters, calibrated to data targets also sampled from uncertainty distributions^4^. We repeated the analysis for each set of parameters and inputs, generating a distribution for each outcome, from which we report summary values (mean or median) and 95% uncertainty intervals.

We additionally performed several sensitivity analyses. First, we examined how variation in within-prison tuberculosis incidence could affect the preferred screening strategy. We generated scenarios with lower prison incidence by varying within-prison transmission rates, representing variation in prison conditions (i.e. crowding, ventilation). We then conducted multinomial logistic regression and estimated the probability that each strategy would be optimal across the range of prison incidence levels. Second, we re-examined the efficiency frontier and preferred strategy in a scenario without access to CXR-CAD technology, which remains the current reality for most prisons in low- and middle-income countries. Third, we assessed how costs and health benefits would vary with reduced screening coverage, focusing on periodic prison-wide screening with CXR-CAD. Additional details are provided in the **Appendix**.

## RESULTS

### Carceral characteristics and status quo tuberculosis incidence

The median projected incarceration rate in 2026 among the population aged 15 and older was 286 per 100,000 in Colombia, 376 per 100,000 in Peru, and 476 per 100,000 in Brazil (**Figure S3**). Carceral dynamics also varied across countries. The average duration of incarceration ranged from 1.3 years in Brazil to 4.9 years in Peru, and the proportion entering prison with prior incarceration history ranged from 18% in Colombia to 60% in Brazil.

In the base-case scenario without systematic screening in prisons, the model projected that by 2035, the incidence of tuberculosis in prisons would be approximately 2000 (95% UI, 1600-2700) per 100,000 person-years in Brazil, 1500 (1000-2500) per 100,000 in Colombia, and 6100 (4100-9600) per 100,000 in Peru (**Figure S3**). At the population level, projected tuberculosis incidence in 2035 without screening in prisons was 33 (95% UI, 28-41) per 100,000 in Colombia, 42 (38-47) per 100,000 in Brazil, and 105 (83-130) per 100,000 in Peru.

### Intervention impacts on prison and population-wide tuberculosis incidence

Annual screening was projected to significantly reduce in-prison and population-wide tuberculosis incidence across countries. Compared to the base-case scenario, annual screening with CXR-CAD could reduce prison incidence in 2035 by 35.3% (95% UI, 27.9-43.8) in Brazil, 58.7% (48.4-69.4) in Colombia, and 32.3% (24.2-39.7) in Peru (**Figure 1, Table S2**). These correspond to population-level incidence reductions of 13.8% (95% UI, 10.2-18.7) in Brazil, 11.9% (7.2-20.7) in Colombia, and 10.6% (7.4-14.2) in Peru.

**Figure 1.**
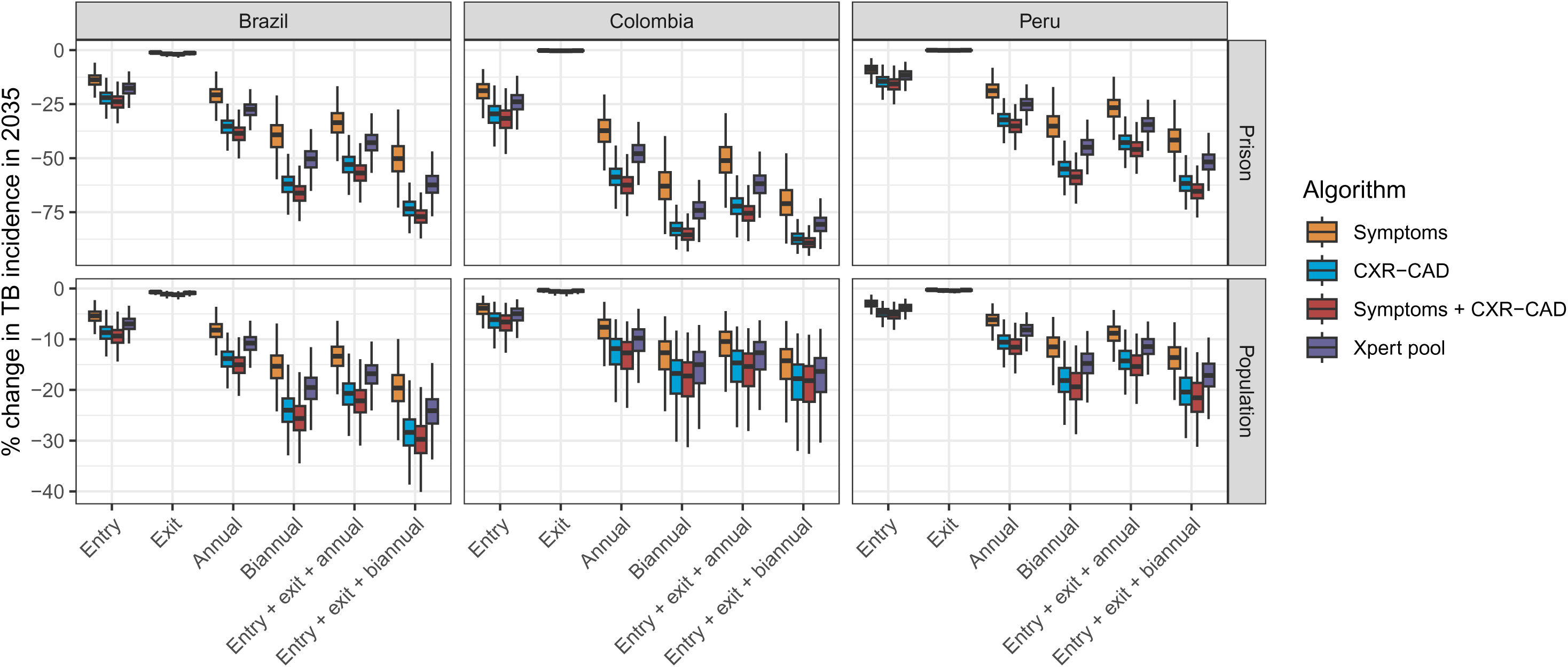
Impacts of screening interventions on within-prison and population-wide tuberculosis incidence in 2035. CXR-CAD, chest X-ray with computer-aided detection.

Biannual (twice-yearly) screening could yield even greater impacts than annual screening. Biannual screening with CXR-CAD was projected to reduce prison incidence in 2035 by 61.9% (95% UI, 52.0-71.9) in Brazil, 83.1% (74.7-89.6) in Colombia, and 55.0% (45.7-63.9) in Peru (**Figure 1, Table S2**). At the population level, biannual screening with CXR-CAD could reduce incidence by 24.0% (95% UI, 18.1-31.2) in Brazil, 16.8% (10.4-28.4) in Colombia, and 18.1% (12.6-24.4) in Peru. Compared to annual screening with CXR-CAD, biannual screening with CXR-CAD averted, on average, 58% more DALYs in Brazil, 35% more DALYs in Colombia, and 46% more DALYs in Peru (**Table 2**). Biannual screening even averted more DALYs than the combination of entry, exit, and annual screening, highlighting the favorable effectiveness of prison-wide screening (**Table S3**).

**Table 2.** Costs, effects, and cost-effectiveness of strategies on the efficiency frontier. Mean estimates are shown, standardized per 100,000 population. All estimates are population-wide (i.e., they include costs and effects accrued by the entire population, not just those in prison). Costs are in 2023 US dollars (USD). Only strategies on the cost-effectiveness efficiency frontier are shown; all strategies are shown in **Table S3**. DALYs, disability-adjusted life years; increm, incremental; ICER, incremental cost-effectiveness ratio.

| Strategy (intervention, algorithm) | Cost (USD) | Effect (DALYs averted) | Increm. cost | Increm. effect | ICER |
| --- | --- | --- | --- | --- | --- |
| **Brazil** |  |  |  |  |  |
| Base case | 304 676 | 0 |  |  |  |
| Annual, CXR-CAD | 342 428 | 132 | 37 752 | 132 | 286 |
| Biannual, CXR-CAD | 385 659 | 208 | 43 231 | 76 | 565 |
| Entry+exit+biannual, CXR-CAD | 500 133 | 247 | 114 474 | 38 | 2984 |
| Entry+exit+biannual, symptoms & CXR-CAD | 660 055 | 257 | 159 922 | 10 | 15416 |
| **Colombia** |  |  |  |  |  |
| Base case | 233 696 | 0 |  |  |  |
| Annual, CXR-CAD | 256 909 | 120 | 23 213 | 120 | 193 |
| Biannual, CXR-CAD | 284 616 | 163 | 27 707 | 42 | 654 |
| Entry+exit+biannual, CXR-CAD | 320 799 | 175 | 36 183 | 12 | 2925 |
| Entry+exit+biannual, symptoms & CXR-CAD | 391 939 | 180 | 71 140 | 4 | 16207 |
| **Peru** |  |  |  |  |  |
| Base case | 1 017 841 | 0 | NA | NA | NA |
| Annual, CXR-CAD | 1 045 096 | 426 | 27 254 | 426 | 64 |
| Biannual, CXR-CAD | 1 060 596 | 621 | 15 501 | 195 | 79 |
| Entry+exit+biannual, CXR-CAD | 1 098 123 | 679 | 37 526 | 58 | 645 |
| Entry+exit+biannual, symptoms & CXR-CAD | 1 203 201 | 704 | 105 078 | 25 | 4158 |

Combining entry, exit, and biannual screening was the most impactful strategy. Conducted with CXR-CAD, this combination was projected to reduce prison incidence by 73.5% (95% UI, 63.6-81.9) in Brazil, 86.5% (82.7-91.7) in Colombia, and 61.4% (52.9-69.6) in Peru (**Figure 1, Table S2**). This amounted to population tuberculosis incidence reductions of 28.4% (95% UI, 21.6-36.5) in Brazil, 17.8% (11.0-29.7) in Colombia, and 20.4% (14.1-27.2) in Peru.

After interventions ended, impacts were not sustained, with incidence returning to baseline levels within five to ten years (**Figure S4**).

### Comparative impacts of different screening algorithms

Algorithms involving CXR-CAD were most impactful in reducing prison and population incidence. When used for biannual screening, CXR-CAD was projected to yield 33-58% greater reductions in prison incidence than symptom screening across countries (**Table S2**). Projected impacts from screening with both symptoms and CXR-CAD were slightly greater than, but largely comparable to, those from screening with CXR-CAD alone (**Figure 1, Table S2**).

Following algorithms involving CXR-CAD, the next most impactful algorithm was GeneXpert with sputum pooling, which yielded 20-28% greater projected reductions in prison incidence than symptom screening when used biannually (**Table S2**).

Projected impacts of symptom screening were less than those of more sensitive algorithms but still substantial, especially with periodic prison-wide screening. In fact, biannual symptom screening had projected impacts comparable to those of annual screening with CXR-CAD (**Table 2, Table S2**).

### Costs and cost-effectiveness of screening strategies

**Table S3** reports DALYs averted, total costs, and additional costs relative to the base case scenario for all interventions and screening algorithms evaluated. Of all screening strategies assessed, four were on the efficiency frontier, exhibiting the greatest health benefit (i.e., averted the most DALYs) at a given cost (**Figure 2**, **Table 2**). All other strategies were strictly dominated (i.e., offer less health benefit at equivalent or higher cost) or weakly (extended) dominated (i.e., an alternative exists that offers better value for money) (**Table S3**).

**Figure 2.**
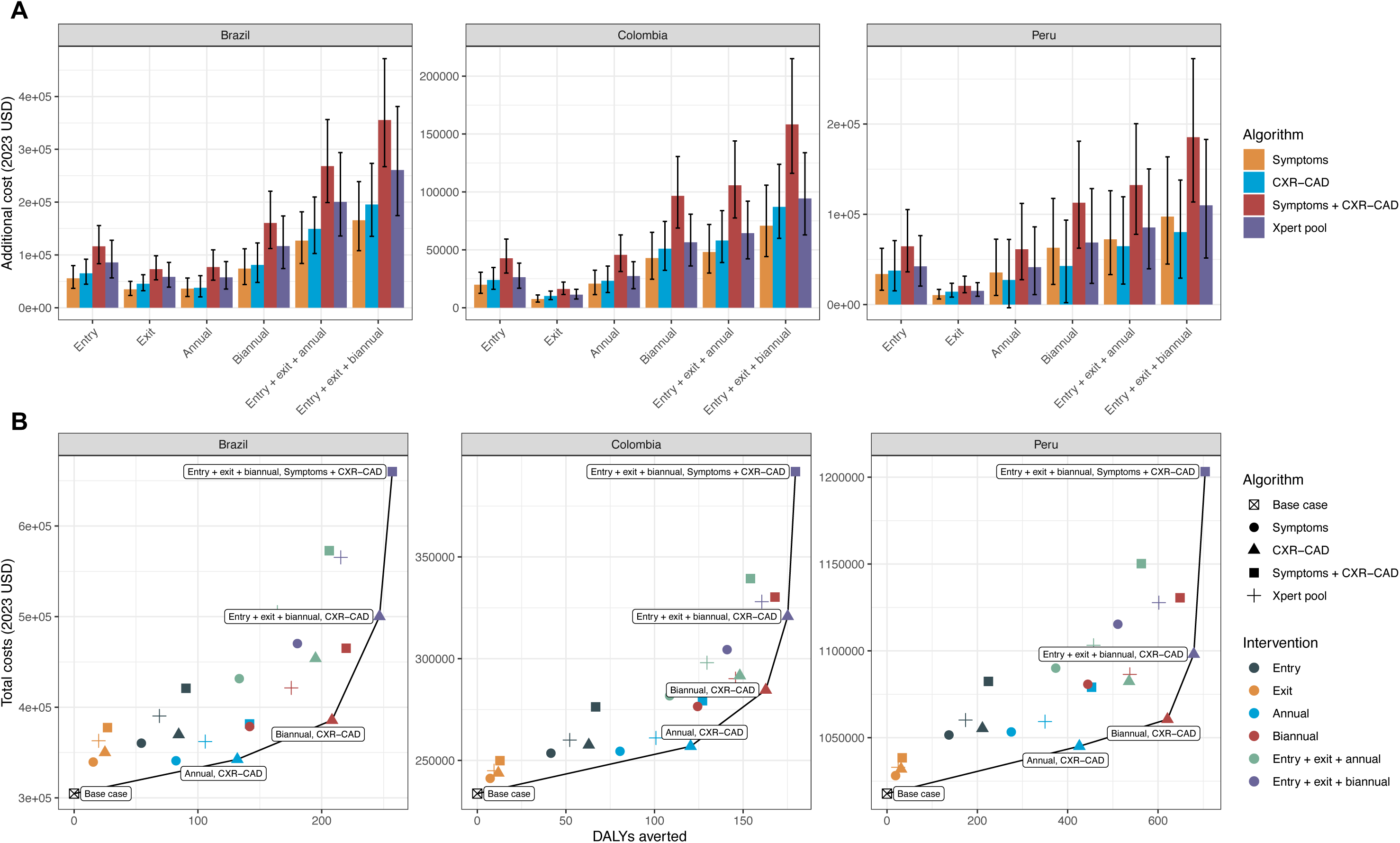
Costs and cost-effectiveness of screening interventions. A) Mean additional cost of screening interventions relative to the base-case scenario. The error bars indicate the 95% uncertainty interval. B) Cost-effectiveness planes for each country. Strategies on the efficiency frontier are highlighted. Costs are in 2023 USD. CXR-CAD, chest x-ray with computer-aided detection; DALYs, disability-adjusted life years.

The four non-dominated strategies on the efficiency frontier, ordered from least to most costly and effective, were:

1. Annual screening with CXR-CAD;
2. Biannual screening with CXR-CAD;
3. Entry, exit, and biannual screening with CXR-CAD; and
4. Entry, exit, and biannual screening with symptoms and CXR-CAD.

In Brazil and Colombia, strategy 3 (entry, exit, and biannual screening with CXR-CAD) was the optimal strategy, providing the greatest health benefit within each country’s cost-effectiveness threshold. Specifically, upgrading from biannual screening to the combination of entry, exit, and biannual screening with CXR-CAD had an ICER of $2984 in Brazil and $2925 in Colombia (**Table 2**). Moving from strategy 3 to 4 (i.e., screening with both symptoms and CXR-CAD rather than CXR-CAD alone) averted few additional DALYs at much higher cost, with ICERs of $15,416 in Brazil and $16,207 in Colombia, exceeding both countries’ cost-effectiveness thresholds.

In Peru, the ICER for moving from strategy 3 to 4 was $4158, within the cost-effectiveness threshold, suggesting that, in the highest burden settings, the additional benefits justify the additional costs of a slightly more sensitive but less specific screening algorithm (**Table 2**).

Of note, the combination of entry, exit, and annual screening—the current WHO recommendation—was dominated by biannual screening in all three countries (**Figure 2, Table S3**). In other words, countries that have implemented annual screening would get more health benefit while spending less by increasing screening frequency to twice yearly, rather than adding entry and exit screening.

### Sensitivity analysis: varying prison incidence

We examined how the optimal strategy would vary by prison incidence, by simulating scenarios with differing levels of within-prison transmission. Entry, exit, and biannual screening with CXR-CAD remained the optimal strategy at prison incidence levels above approximately 1,000 per 100,000 person-years (**Figure 3, Table S4**). At extraordinarily high prison incidence levels (>5,300 per 100,000 person-years), combining symptoms and CXR-CAD for entry, exit, and biannual screening became the preferred strategy.

**Figure 3.**
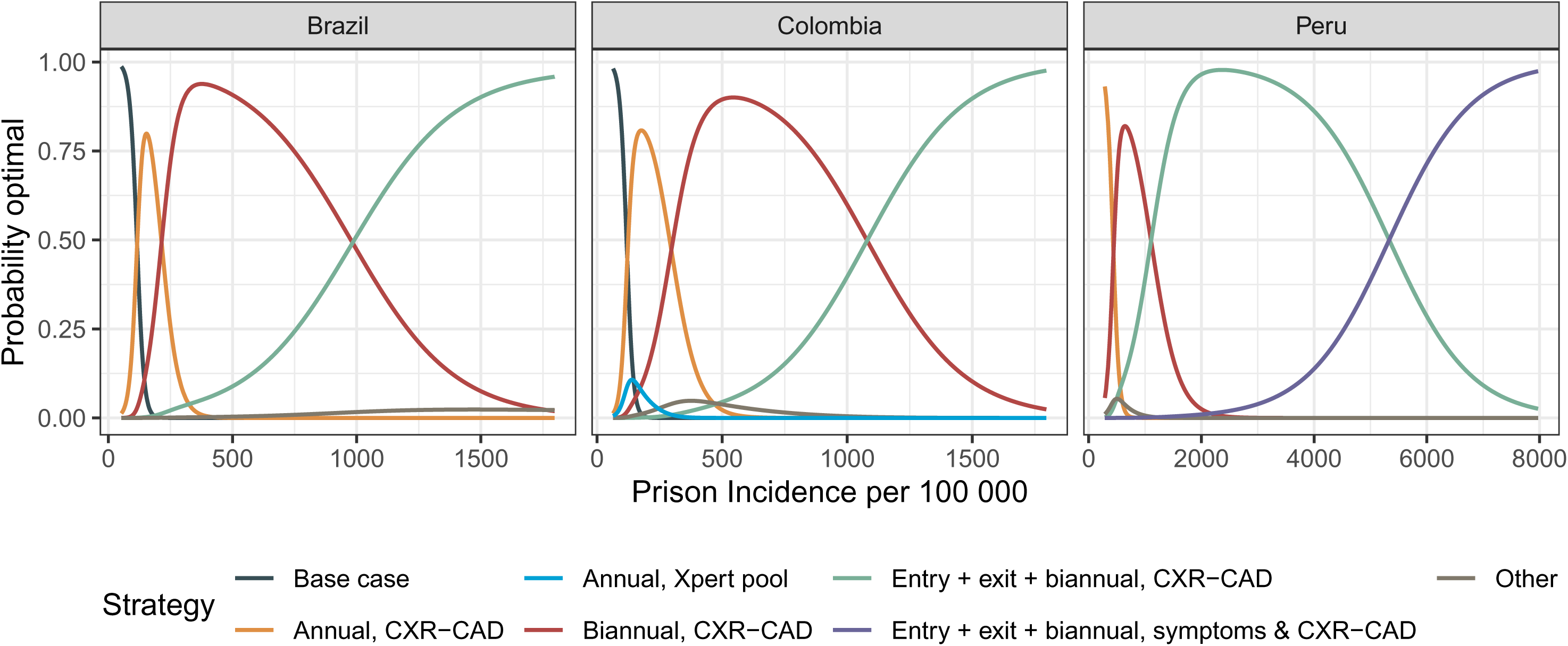
Optimal strategies under varying prison incidence scenarios. Prison incidence was varied by varying the rate of within-prison transmission. The probability that a given strategy was optimal across a range of prison incidence was modeled using multinomial logistic regression. Only strategies with modeled probabilities greater than or equal to 5% in at least one incidence quartile are shown; all other strategies are lumped into “other”. Prison incidence under approximately 300 per 100,000 was not modeled in Peru due to parameter constraints (see Appendix, sensitivity analyses).

At lower prison incidence levels, less intensive strategies were optimal. Annual screening with CXR-CAD was cost-effective at prison incidence levels greater than approximately 120 per 100,000 person-years (**Figure 3, Table S4**). Biannual screening with CXR-CAD was cost-effective at prison incidence levels above approximately 210 per 100,000 in Brazil, 300 per 100,000 in Colombia, and 440 per 100,000 in Peru.

### Sensitivity analysis: no CXR-CAD

If CXR-CAD were unavailable, the best alternative strategy would be entry, exit, and biannual screening using Xpert on pooled sputum (**Table S5**). Compared to the next best strategy (biannual screening with pooled Xpert), the combination of entry, exit, and biannual screening with pooled Xpert had an ICER of $3607 in Brazil, $2551 in Colombia, and $642 in Peru. Other strategies on the efficiency frontier included annual and biannual screening with symptoms or pooled Xpert, depending on the country (**Figure S7**).

## DISCUSSION

Evidence is lacking to guide optimal strategies for tuberculosis active case-finding in prisons. In this study, we used a dynamic transmission model to project the population-wide impacts and cost-effectiveness of different prison-based screening approaches in Brazil, Colombia, and Peru. We find that combined entry, exit, and biannual screening with CXR-CAD is the most impactful and cost-effective strategy in Brazil and Colombia, while a more sensitive algorithm (combining symptoms and CXR-CAD) is justified in Peru’s higher-burden prisons. These results support prioritizing intensive, periodic prison-based screening up to twice yearly, particularly in settings where prisons drive tuberculosis transmission.

The WHO recommends prison-wide screening at least annually and has stated that biannual mass screening, while “ideal,” may not be sustainable due to cost and logistical barriers^6,11^. As the first modeling study to evaluate screening more than once a year, we found that biannual prison-wide screening, alone or in combination with entry and exit screening, was highly cost-effective across heterogeneous countries and led to substantial reductions in tuberculosis incidence in prisons and in the general population. Importantly, these intensive strategies remained cost-effective at prison incidence levels far below national averages, with biannual CXR-CAD alone remaining cost-effective at levels as low as 200–400 per 100,000. These findings strongly support implementation and regional scale-up of twice-yearly screening in prisons, with investments justified by their projected population-wide benefits.

Regarding screening algorithms, strategies involving CXR-CAD were consistently the most impactful and cost-effective, aligning with empirical studies demonstrating its feasibility and cost-efficiency in prison settings^7,8,20–22^. Prior modeling work in other regions has also supported the value of CXR-based screening in prisons^13,23^. In Brazil and Colombia, entry, exit, and biannual screening with CXR-CAD alone was optimal. In Peru, where the model projected prison incidence to exceed 5000 cases per 100,000 person-years without screening, combining symptoms and CXR-CAD was cost-effective, reflecting a shift toward prioritizing sensitivity over specificity. Of note, in 2023 the Peru National Tuberculosis Program (DPCTB) began implementing entry and annual screening using this same algorithm, reaching nearly 40,000 PDL across 18 prisons^8^. Overall, our results strongly support using CXR-CAD for systematic screening in prisons. While implementation requires upfront investments in equipment, software, and training, we accounted for these costs in our analysis, and CXR-CAD remained the cost-effective algorithm. Moreover, advances in ultra-portable X-ray technology may further improve scalability and sustainability of intensive periodic screening by enabling coverage of multiple prisons with a single device^10^.

In the absence of CXR-CAD, pooled Xpert was a favorable alternative, averting 87-92% as many DALYs as CXR-CAD and remaining highly cost-effective across countries. While most symptom-based strategies were dominated, intensive (i.e. biannual) symptom screening nonetheless averted substantial DALYs—more than annual screening with CXR-CAD. These findings highlight the importance of immediately implementing prison-wide screening with available tools while developing capacity for more sensitive algorithms.

Entry or exit screening alone had modest impacts and should be performed in addition to periodic prison-wide screening. Entry screening may integrate smoothly into prison intake procedures but has lower yield per person screened due to the lower tuberculosis prevalence at entry^12,24^. Exit screening also showed limited impact and may present substantial logistical challenges under routine programmatic conditions. Nonetheless, given the high risk of tuberculosis and barriers to continuity of care post-release, it remains a strategy worth exploring in future work.

Notably, we project that if any screening intervention were to end without additional efforts to reduce within-prison transmission, tuberculosis rates would rebound rapidly to pre-intervention levels. To enable sustained progress toward tuberculosis elimination, intensive screening should be paired with complementary interventions like tuberculosis preventive therapy, as well as structural efforts to improve prison conditions and reduce incarceration rates^4,25,26^.

There are several limitations to note. Our model did not include age, gender, or HIV, although HIV prevalence is relatively low in these countries^27^. Our simplified approach to representing MDR-TB does not capture complexities of longer treatment regimens or diagnostic delays, which may influence preferred screening strategies. A study modeling tuberculosis screening in prisons in the former Soviet Union, where MDR-TB rates are high, found that annual screening with Xpert was more effective in reducing MDR-TB prevalence compared to annual radiographic screening^23^. Pooled Xpert may be a useful tool in high MDR-TB prisons but should be tested empirically. Furthermore, we did not explicitly model prison transfers, which could reduce effectiveness if screening is not coordinated across facilities. This may partly explain the discrepancy between our results and a recent empirical study in Brazil, which found no reduction in tuberculosis prevalence after three rounds of prison mass screening between 2017 and 2021^28^. In that study, screening was limited to three of 29 male prisons in the state, and frequent transfers likely diluted impact. Future work might examine how inter-facility movement and variation in screening practices across facilities affect intervention effectiveness. Finally, we did not model heterogeneity in infectiousness. If a small fraction of individuals accounts for most transmission, imperfect screening coverage may allow continued transmission. Collectively, these factors highlight the complexities of real-world implementation and the need for empirical studies to validate and refine model projections and evaluate feasible strategies for operationalization under programmatic conditions.

Altogether, our findings support the prioritization of combined entry, exit, and biannual screening with CXR-CAD in Brazil, Colombia, and Peru, and similar settings. This strategy is projected to be highly cost-effective and impactful in reducing tuberculosis morbidity and transmission in prisons and population-wide. Screening should begin immediately with available methods while infrastructure is developed for more impactful and cost-effective tools like CXR-CAD. Ultimately, sustained investments in intensive, prison-wide screening—alongside structural reforms to reduce incarceration and prison crowding—will be essential to advancing regionally toward tuberculosis elimination.

## Data Availability

All data produced in the present work are contained in the manuscript and supplementary files.

