## Supplementary Appendix for "Comparative impacts and cost-effectiveness of tuberculosis active case-finding strategies in prisons in Brazil, Colombia, and Peru: a mathematical modeling study"

**Table of Contents**

**Methodological Details**

**Supplementary Tables**

Table S1. Additional parameters and cost inputs for base-case scenario and screening interventions.

Table S2. Impacts of screening interventions on prison and population TB incidence in 2035.

Table S3. Health benefits and costs of screening interventions.

Table S4. Optimal strategies by prison incidence.

Table S5. Costs, effects, and cost-effectiveness of strategies on the efficiency frontier if CXR-CAD were unavailable.

**Supplementary Figures**

Figure S1. Model fit to incarceration-related data targets.

Figure S2. Model fit to tuberculosis-related data targets.

Figure S3. Carceral characteristics and projected tuberculosis incidence in included countries.

Figure S4. Tuberculosis incidence over time under base-case and intervention scenarios.

Figure S5. Total costs of base-case scenario and screening interventions.

Figure S6. Additional costs of screening interventions, relative to base-case scenario, disaggregated by costs of passive diagnosis, testing, and treatment.

Figure S7. Cost-effectiveness plane without algorithms using CXR-CAD.

**Methodological Details**

*Calibration targets*

Calibration targets for within-prison tuberculosis incidence were based on a recent study using Bayesian meta-regression modeling to generate national and regional estimates of prison incidence and the case detection ratio (CDR)^1^. For Brazil and Colombia, where country-specific data were included from active-case finding studies in prisons, we directly used posterior distributions of prison incidence from this study. For Peru, where no such active case-finding studies had been conducted (prior to 2023), we estimated the uncertainty distribution for the prison incidence calibration target by applying a regional CDR of 0.54 (95% UI, 0.20-0.96) to Peru-specific prison notifications data.

*Probability of treatment success during and after incarceration*

To determine probability of treatment success, we consider probability of treatment completion as well as culture conversion among patients who do not complete the full treatment regimen, assuming that one third of patients who do not complete treatment are nonetheless cured^2^. A recent analysis in Mato Grosso do Sul, Brazil found that 67% of individuals newly diagnosed with tuberculosis during incarceration complete treatment^3^. Accounting for cure without full treatment completion results in a 78% probability of treatment success for individuals diagnosed during incarceration. The same analysis found that 61% of individuals diagnosed within two years post-release complete treatment, which we assume to be comparable for individuals diagnosed through exit screening. Accounting for possible cure without full treatment completion results in 74% treatment success for individuals diagnosed through exit screening.

*Impacts of COVID-19 pandemic*

Prior assumptions about the impact of the COVID-19 pandemic on tuberculosis were updated based on newly available data. Specifically, we assumed that tuberculosis diagnosis rates were less impacted in prisons than in the community^4^, and that return to pre-COVID-19 pandemic diagnosis rates in all strata would occur gradually over several years.

*Passive diagnosis*

To estimate usage of various diagnostic methods under the base-case scenario, we use data from the WHO Global TB Report (and for Brazil, the Information System for Notifiable Diseases SINAN). We assume that patients undergo clinical evaluation and receive any combination of smear, molecular rapid diagnostics (i.e. GeneXpert), culture, and drug sensitivity testing, with the proportion of patients receiving each assay sampled from independent distributions.

To estimate costs of passive diagnosis and treatment, we estimated, per person cured: 1) the number of people evaluated for tuberculosis, 2) the number of people diagnosed with tuberculosis, and 3) the number of people initiated on treatment. According to data from the WHO Global TB Report 2023 for Brazil, 12% of individuals initially tested with a WHO-recommended rapid diagnostic test (WRD) had a positive test result, and 6.2 WRDs were used per person notified as a tuberculosis case. Since WRDs are not used for everyone who is evaluated (45% of people newly diagnosed with tuberculosis in Brazil in 2022 were initially tested with a WRD), the number of individuals evaluated per person diagnosed is likely higher. We therefore assume approximately 10 (range 5-15) individuals are evaluated for tuberculosis per person diagnosed. To adjust this ratio to be a function of the number of people evaluated per person cured, we use a recent study on the tuberculosis care cascade in Brazil which showed that among individuals diagnosed with tuberculosis, 90% initiate treatment and 81.4% are cured^5^. We assume the same ratios across countries.

*Test accuracy*

For each screening or diagnostic method (cough, CXR-CAD, Xpert Ultra), we simulated joint uncertainty distributions for sensitivity and specificity, with induced correlation of -0.5 to account for their inverse dependency.

*Costs*

Country-specific costs were not available for Colombia and, in some cases, Peru. For these costs, we borrowed parameters from Brazil, assuming the same cost of consumables and applying an adjustment factor for labor costs based on ratios of GDP per capita.

*Cost-effectiveness analysis*

To project lifetime years of life lost (YLLs) for people who die from tuberculosis, we calculated the average age of people developing tuberculosis among the population 15 and older^6^ (37 in Brazil, 40 in Colombia, and 37 in Peru) and retrieved remaining life expectancy at that age (**Table S1**). We assumed similar life expectancy across population strata. To project years lived with disability (YLDs) due to post-tuberculosis sequelae, we applied a post-tuberculosis disability weight over the remaining life expectancy of individuals who recover from tuberculosis (**Table S1)**. At the end of the analytic horizon, individuals with tuberculosis were assumed to either recover or die immediately from tuberculosis with probabilities based on competing risks. We did not include elevated all-cause mortality for tuberculosis survivors given uncertainty around the true causal risk ratio.

Cost-effectiveness thresholds were drawn from recent economic evaluations commissioned by ministries of health in each of the selected countries and were similar to the crude estimate of GDP per capita often used for low- and middle-income countries^7-9^. Thresholds were updated for 2023 using time trends in GDP per capita. We did not project future healthcare costs beyond the ten-year analytic horizon, nor account for future infection, disease, or transmission occurring after the end of this period.

*Sensitivity analyses*

For the first sensitivity analysis, we generated a wide range of within-prison tuberculosis incidence by multiplying the previously calibrated effective contact rate in prison by a sampled factor 0.1 < *f* < 1. A constraint was then imposed to ensure that the prison effective contact rate in prison remained greater than or equal to the community effective contact rate. After re-simulating screening interventions, we identified the optimal strategy, defined as the most effective, non-dominated strategy with an incremental cost-effectiveness ratio (ICER) within the country’s cost-effectiveness threshold. We then fit a multinomial logistic regression model with the optimal strategy as the outcome and within-prison incidence as the predictor, and we used this model to estimate the probability that each strategy would be optimal across the incidence range.

Each sensitivity analysis was conducted with 1000 sets of model parameters and inputs, combining uncertainty from existing model parameters and inputs with additional sampled parameters for the sensitivity variable(s) of interest. This ensured that uncertainty was propagated throughout the sensitivity analyses.

**Table S1. Additional parameters and cost inputs for base-case scenario and screening interventions.** All parameters were sampled from triangle distributions, for which the mode (range) is listed. All costs are in 2023 US dollars (USD). Note: costs of methods used within <Note that costs here are for individual things not entire algorithm (ie CXR-CAD screening also includes symptom interview)

| Parameter | Country | Value (Range) | Source |
| --- | --- | --- | --- |
| Base case scenario (passive diagnosis) | | | |
| Proportion evaluated with GeneXpert | Brazil | 0.44 (0.4-0.5) | ^6,10^ |
|  | Colombia | 0.53 (0.45-0.65) | ^1^ |
|  | Peru | 0.3 (0.25-0.4) | ^1^ |
| Proportion evaluated with smear | Brazil | 0.7 (0.6-0.8) | ^6,10^ |
|  | Colombia | 0.81 (0.7-0.9) | ^1^ |
|  | Peru | 0.8 (0.7-0.9) | ^1^ |
| Proportion of patients receiving culture | Brazil | 0.45 (0.4-0.5) | ^6,10^ |
|  | Colombia | 0.42 (0.35-0.5) | ^1^ |
|  | Peru | 0.62 (0.5-0.75) | ^1^ |
| Proportion of patients receiving drug susceptibility testing | Brazil | 0.25 (0.1-0.33) | ^6,10^ |
|  | Colombia | 0.6 (0.4-0.7) | ^1^ |
|  | Peru | 0.69 (0.6-0.8) | ^1^ |
| Average number of people evaluated per person cured | All | 12.29 (6.14-18.43) | ^5,6^ |
| Average number of people diagnosed per person cured | All | 1.23 (1.17-1.31) | ^5^ |
| Average number of people initiated on treatment per person cured | All | 1.11 (1.05-1.2) | ^5^ |
| Screening interventions | | | |
| Screening coverage | All | 0.8 (0.7-0.9) | ^11-13^ |
| Duration of screening round for periodic screening (years) | All | 0.167 (0.083-0.25) | Assumed |
| Probability of DS-TB treatment completion in prison for individuals detected through ACF | All | 0.67 (0.6-0.75) | ^3^ |
| Probability of linkage to care post-release for individuals detected through exit screening | All | 0.65 (0.5-0.75) | Assumed |
| Probability of DS-TB treatment completion post-release following linkage to care | All | 0.61 (0.5-0.7) | Unpublished data from Brazil |
| Proportion of TB patients with RR-TB, general population | Brazil | 0.016 (0.006-0.025) | ^6^ |
|  | Colombia | 0.045 (0.02-0.06) | ^6^ |
|  | Peru | 0.0726 (0.06-0.085) | ^14^ |
| Proportion of TB patients with RR-TB, prison | Brazil | 0.016 (0.006-0.025) | Assumed same as general population |
|  | Colombia | 0.045 (0.02-0.06) | Assumed same as general population |
|  | Peru | 0.093 (0.08-0.12) | ^14^ |
| Probability of detection and treatment completion for individuals with RR-TB detected through ACF, relative to individuals with DS-TB | All | 0.4875 (0.4-0.6) | ^6^ |
| Cost-effectiveness analysis | | | |
| Disability weight for active TB disease | All | 0.33 (0.25-0.4) | ^15^ |
| Post-TB disability weight | All | 0.038 (0.02-0.05) | ^16^ |
| Remaining life expectancy at average age of developing TB among population aged 15 and older (years) | Brazil | 44 (39-48) | ^17^ |
|  | Colombia | 42 (37-47) | ^17^ |
|  | Peru | 47 (42-52) | ^17^ |
| Per-unit cost of smear microscopy | Brazil | 3.9 (2-6) | ^18^ |
|  | Colombia | 3.01 (1.54-4.63) | ^18^ |
|  | Peru | 2.14 (1.5-5) | ^19^ |
| Per-unit cost of culture | Brazil | 10.43 (8-22) | ^18^ |
|  | Colombia | 8.84 (6.17-16.97) | ^18^ |
|  | Peru | 9.26 (7.1-19.53) | ^18^ |
| Per-unit cost of drug susceptibility testing (assuming 50/50 LPA vs. culture-based testing) | Brazil | 111.1 (55-165) | ^20^ |
|  | Colombia | 94.15 (42.42-127.25) | ^20^ |
|  | Peru | 154.49 (77.24-231.73) | ^21^ |
| Per-unit cost of Xpert MTB/RIF Ultra, accounting for recent cartridge price reduction | Brazil | 18.99 (17-35) | ^11^ |
|  | Colombia | 16.86 (14.25-26.75) | ^11^ |
|  | Peru | 17.42 (14.97-28.93) | ^11^ |
| Per-unit additional cost of pooled Xpert (50% additional human resources) | Brazil | 1.72 (1.5-2) | ^11^ |
|  | Colombia | 1.20 (1.04-1.39) | ^11^ |
|  | Peru | 1.33 (1.16-1.55) | ^11^ |
| Per-unit cost of CXR-CAD | Brazil | 7.5 (6-11) | ^11^ |
|  | Colombia | 6.83 (5.8-10) | ^11^ |
|  | Peru | 7.01 (5.5-10) | ^11^ |
| Per-unit cost of symptom interview | Brazil | 2.27 (2-3) | ^11^ |
|  | Colombia | 1.58 (1.39-2.08) | ^11^ |
|  | Peru | 1.76 (1.5-2.5) | ^11^ |
| Per-unit cost of clinical evaluation | Brazil | 3.11 (2-5) | ^11^ |
|  | Colombia | 2.16 (1.8-5) | ^11^ |
|  | Peru | 2.41 (2-6) | ^11^ |
| Cost of drug-sensitive treatment | Brazil | 567 (460-800) | ^20^ |
|  | Colombia | 497.81 (403.87-702.38) | ^20^ |
|  | Peru | 540.71 (440-800) | ^21^ |
| Cost of RR-TB treatment (assuming 50/50 standard 18-month regimen vs. BPaLM) | Brazil | 5301.5 (2650-8000) | ^20^ |
|  | Colombia | 4816.32 (2407.48-7267.86) | ^20^ |
|  | Peru | 5098.15 (3000-8000) | ^21^ |
| Relative cost of incomplete treatment | All | 0.5 | Assumed |

**Table S2. Impacts of screening interventions on prison and population tuberculosis incidence in 2035.** Median estimates and 95% uncertainty intervals are shown for the projected percent reduction in prison or population incidence in 2035, relative to the base-case scenario.

| Country | Intervention | Population | Symptoms | CXR-CAD | Symptoms + CXR-CAD | Pooled Xpert |
| --- | --- | --- | --- | --- | --- | --- |
| Brazil | Entry | Prison | 13.7 (8.6-21.1) | 22 (15.3-30.4) | 23.8 (16.8-32.4) | 17.6 (12.1-24.9) |
|  |  | Population | 5.4 (3.2-8.6) | 8.7 (5.8-12.9) | 9.4 (6.3-13.9) | 7 (4.6-10.5) |
|  | Exit | Prison | 1.1 (0.5-2) | 1.7 (0.9-3) | 1.9 (1-3.4) | 1.4 (0.7-2.4) |
|  |  | Population | 0.7 (0.4-1.2) | 1.1 (0.7-1.8) | 1.2 (0.8-2) | 0.9 (0.5-1.5) |
|  | Annual | Prison | 20.7 (13.6-30.5) | 35.3 (27.9-43.8) | 38.5 (30.6-47.3) | 27.4 (21.2-35.2) |
|  |  | Population | 8.2 (5.1-12.7) | 13.8 (10.2-18.7) | 15 (11.2-20.1) | 10.8 (7.8-15) |
|  | Biannual | Prison | 39.2 (26.4-54.7) | 61.9 (52-71.9) | 66.2 (56.2-75.6) | 50.3 (40.6-61.7) |
|  |  | Population | 15.3 (9.8-22.6) | 24 (18.1-31.2) | 25.6 (19.5-33) | 19.5 (14.4-26.2) |
|  | Entry + exit + annual | Prison | 33.6 (22.4-47.3) | 53 (42.8-63.5) | 56.9 (46.8-66.9) | 42.8 (33.1-53.6) |
|  |  | Population | 13.3 (8.3-19.7) | 20.7 (15.3-27.7) | 22.1 (16.7-29.4) | 16.8 (12.2-23.3) |
|  | Entry + exit + biannual | Prison | 50.2 (34.4-66.9) | 73.5 (63.6-81.9) | 77.1 (67.9-84.4) | 62.4 (51.2-73.1) |
|  |  | Population | 19.6 (12.7-28.2) | 28.4 (21.6-36.5) | 29.7 (22.7-38) | 24.1 (18-32) |
| Colombia | Entry | Prison | 19.3 (10.3-25.5) | 28.3 (21.8-39.8) | 30.6 (24.3-42.8) | 23 (17-32.3) |
|  |  | Population | 3.3 (2.3-6.6) | 5.5 (4.4-10.4) | 5.8 (4.8-11.1) | 4.5 (3.6-8.5) |
|  | Exit | Prison | 0.1 (0.1-0.3) | 0.2 (0.1-0.5) | 0.3 (0.2-0.5) | 0.2 (0.1-0.4) |
|  |  | Population | 0.3 (0.1-0.6) | 0.5 (0.3-1) | 0.5 (0.3-1.1) | 0.4 (0.2-0.8) |
|  | Annual | Prison | 37.5 (23.2-48.4) | 57.6 (50.1-66.4) | 62.7 (54.8-70.3) | 48.1 (37-56) |
|  |  | Population | 6.8 (4.4-11.2) | 10.7 (8.9-17.4) | 11.6 (9.6-18.4) | 8.5 (6.9-14.4) |
|  | Biannual | Prison | 62.1 (42.6-75.2) | 82.8 (76.2-88.1) | 85.2 (80.1-90) | 74.4 (62.3-81.4) |
|  |  | Population | 11.3 (8-17.8) | 15.6 (12.8-23) | 16.3 (13.2-23.7) | 13.3 (11.4-21) |
|  | Entry + exit + annual | Prison | 51.3 (33.8-62.4) | 69.9 (65.6-80.5) | 73.7 (69.7-83.3) | 61 (52-71.5) |
|  |  | Population | 9.1 (6.6-15.5) | 13.7 (11.2-21.1) | 14.5 (11.8-21.9) | 11.2 (9.5-18.7) |
|  | Entry + exit + biannual | Prison | 70.4 (51.5-81.5) | 86.5 (82.7-91.7) | 88.9 (85.5-92.9) | 80.2 (71.2-87) |
|  |  | Population | 12.7 (9.7-20.2) | 16.9 (13.6-24) | 17.4 (13.8-24.5) | 14.9 (12.6-22.7) |
| Peru | Entry | Prison | 8.9 (4.7-14) | 14.1 (8.8-20.8) | 15.2 (9.6-22.1) | 11.4 (7-16.7) |
|  |  | Population | 2.9 (1.7-4.7) | 4.6 (3-6.7) | 5 (3.3-7.2) | 3.7 (2.4-5.5) |
|  | Exit | Prison | 0.1 (0-0.1) | 0.1 (0-0.2) | 0.1 (0-0.2) | 0.1 (0-0.2) |
|  |  | Population | 0.2 (0.1-0.6) | 0.4 (0.2-0.9) | 0.4 (0.2-1) | 0.3 (0.1-0.8) |
|  | Annual | Prison | 19.1 (13-28.1) | 32 (24.5-38.9) | 34.9 (26.8-41.3) | 25.3 (19.1-32) |
|  |  | Population | 6.2 (4.1-9.8) | 10.7 (7.8-13.8) | 11.8 (8.3-14.8) | 8.3 (6.2-11.2) |
|  | Biannual | Prison | 35.3 (25.3-48.6) | 55 (46.4-62.4) | 58.2 (50.4-66) | 45.3 (38.2-53.7) |
|  |  | Population | 11.8 (7.8-18) | 18.7 (12.8-24.6) | 20 (13.7-26.2) | 15.1 (10.5-20.9) |
|  | Entry + exit + annual | Prison | 27.1 (18.1-37.7) | 42.3 (34.8-50) | 45.5 (37.1-53) | 34.2 (27.7-42.3) |
|  |  | Population | 9 (6-13.7) | 14.3 (10.2-19) | 15.5 (11-20.1) | 11.6 (8.1-15.8) |
|  | Entry + exit + biannual | Prison | 42 (29.9-54.9) | 61.4 (52.9-69.6) | 64.6 (56.7-72.8) | 52 (43.4-60.5) |
|  |  | Population | 14 (9.6-21) | 20.6 (14.6-27.4) | 21.9 (15.4-28.8) | 17.4 (12-24) |

**Table S3. Health benefits and costs of screening interventions.** Median estimates and 95% uncertainty intervals for disability-adjusted life years (DALYs) averted, total costs, and additional costs relative to the base-case scenario over the ten-year intervention period. All estimates are standardized per 100,000 population. Costs are in 2023 US dollars. The “status” column indicates whether a strategy is on the cost-efficient frontier or dominated through strict dominance (D) or extended dominance (ED).

| Intervention | Algorithm | DALYs averted | Total costs | Additional costs relative to base case scenario | Status |
| --- | --- | --- | --- | --- | --- |
| Brazil | | | | |  |
| Base case | N/A | 0 | 304676 (244051-371439) | 0 | Frontier |
| Entry | Symptoms | 54.3 (23.9-112.5) | 360330 (291465-433634) | 55654 (36685-79699) | D |
|  | CXR-CAD | 84.5 (40.9-166.3) | 369771 (304744-439093) | 65095 (44548-91997) | D |
|  | Symptoms + CXR-CAD | 90.4 (43.9-176.7) | 420869 (344571-501427) | 116193 (83416-155554) | D |
|  | Pooled Xpert | 68.9 (32.6-137.9) | 390365 (318286-473837) | 85689 (56425-127858) | D |
| Exit | Symptoms | 15.4 (6-35.6) | 339512 (273935-408911) | 34836 (23244-49967) | ED |
|  | CXR-CAD | 24.9 (10.8-56.9) | 350021 (285794-419018) | 45344 (32170-62447) | D |
|  | Symptoms + CXR-CAD | 26.9 (11.7-62.2) | 377543 (306376-452778) | 72867 (52846-98408) | D |
|  | Pooled Xpert | 19.8 (8.4-44.7) | 362944 (292595-440935) | 58268 (38856-85934) | D |
| Annual | Symptoms | 82.3 (37.6-160.2) | 340920 (275923-408570) | 36244 (21106-56101) | ED |
|  | CXR-CAD | 131.8 (65.2-252.4) | 342428 (281164-406959) | 37752 (20646-60792) | Frontier |
|  | Symptoms + CXR-CAD | 141.7 (70.9-270.2) | 381612 (315921-451025) | 76936 (52247-109496) | D |
|  | Pooled Xpert | 105.9 (51.3-204) | 362100 (295456-435139) | 57424 (35420-87493) | D |
| Biannual | Symptoms | 141.8 (67.8-274.5) | 378660 (307448-451917) | 73984 (43838-111676) | ED |
|  | CXR-CAD | 208.3 (108.2-390.7) | 385659 (320256-452288) | 80983 (47912-122695) | Frontier |
|  | Symptoms + CXR-CAD | 219.7 (114.6-413) | 465089 (385809-548312) | 160413 (112005-220748) | ED |
|  | Pooled Xpert | 175.4 (87.8-332.4) | 421364 (344048-511423) | 116688 (74219-173785) | D |
| Entry + exit + annual | Symptoms | 133.4 (62.7-266) | 431555 (349463-522585) | 126879 (83816-181724) | D |
|  | CXR-CAD | 195 (99.3-373.2) | 453859 (375841-536257) | 149183 (102741-209703) | D |
|  | Symptoms + CXR-CAD | 206.1 (105.7-395.2) | 572795 (465338-694066) | 268119 (199214-356436) | D |
|  | Pooled Xpert | 164.2 (82.2-317.4) | 504948 (409779-626590) | 200272 (135848-293726) | D |
| Entry + exit + biannual | Symptoms | 180.3 (88.6-350) | 470270 (379962-573312) | 165594 (108120-238890) | D |
|  | CXR-CAD | 246.6 (128.3-464.3) | 500133 (413474-597984) | 195457 (135096-273287) | Frontier |
|  | Symptoms + CXR-CAD | 257 (135.5-482.5) | 660055 (537702-799550) | 355379 (267081-471627) | Frontier |
|  | Pooled Xpert | 215.3 (110.9-413.5) | 565428 (453874-713538) | 260752 (174496-381190) | D |
| Colombia | | | | |  |
| Base case | N/A | 0 | 233696 (185309-291875) | 0 | Frontier |
| Entry | Symptoms | 41.6 (12.9-105.2) | 253556 (203136-313984) | 19860 (12391-30622) | ED |
|  | CXR-CAD | 62.9 (20.9-156.6) | 257622 (207849-318121) | 23927 (15918-34728) | D |
|  | Symptoms + CXR-CAD | 66.8 (22.2-164.4) | 276311 (223881-336265) | 42616 (29882-59324) | D |
|  | Pooled Xpert | 52.2 (16.9-130.8) | 260030 (211149-320463) | 26334 (16820-38502) | D |
| Exit | Symptoms | 7.3 (1.8-21.6) | 241179 (191528-300764) | 7484 (4877-10969) | ED |
|  | CXR-CAD | 11.9 (3.2-33.7) | 243830 (194699-303375) | 10135 (6959-14424) | ED |
|  | Symptoms + CXR-CAD | 12.8 (3.5-35.7) | 249877 (199637-312008) | 16182 (11438-22144) | ED |
|  | Pooled Xpert | 9.5 (2.5-26.7) | 244941 (195721-303224) | 11246 (7507-15917) | D |
| Annual | Symptoms | 80.5 (26-194.5) | 254494 (205520-314281) | 20799 (11315-32395) | ED |
|  | CXR-CAD | 120.4 (42.1-288.6) | 256909 (209879-314280) | 23213 (13164-35929) | Frontier |
|  | Symptoms + CXR-CAD | 127 (44.4-300.5) | 279322 (230382-340202) | 45627 (31214-62910) | ED |
|  | Pooled Xpert | 100.8 (34.2-240.6) | 261038 (213087-319393) | 27343 (16432-39704) | D |
| Biannual | Symptoms | 124.3 (42.5-296.1) | 276515 (226881-339450) | 42819 (24600-65079) | ED |
|  | CXR-CAD | 162.8 (59.2-379.4) | 284616 (235236-344845) | 50920 (32287-74515) | Frontier |
|  | Symptoms + CXR-CAD | 168 (61.5-387.6) | 330300 (273776-396095) | 96605 (68795-130613) | D |
|  | Pooled Xpert | 145.7 (52.5-342.3) | 290181 (239029-351242) | 56486 (36091-80727) | D |
| Entry + exit + annual | Symptoms | 108.5 (36.8-259.8) | 281768 (230352-344882) | 48072 (29913-71828) | D |
|  | CXR-CAD | 148.1 (53.2-347.3) | 291655 (240461-352487) | 57959 (39010-83847) | D |
|  | Symptoms + CXR-CAD | 154.1 (55.5-357.1) | 339368 (280034-409019) | 105673 (77467-144019) | D |
|  | Pooled Xpert | 129.6 (46-305.7) | 298039 (243368-361263) | 64343 (42235-92000) | D |
| Entry + exit + biannual | Symptoms | 140.9 (49.4-325.9) | 304457 (248060-370646) | 70761 (44186-105811) | D |
|  | CXR-CAD | 175.1 (64.2-404.2) | 320799 (265685-387393) | 87104 (59913-123801) | Frontier |
|  | Symptoms + CXR-CAD | 179.5 (66.4-413) | 391939 (324151-470299) | 158243 (116039-215054) | Frontier |
|  | Pooled Xpert | 160.5 (58.1-378.4) | 328027 (268947-395029) | 94331 (62884-133815) | D |
| Peru | | | | |  |
| Base case | N/A | 0 | 1017841 (791234-1299987) | 0 | Frontier |
| Entry | Symptoms | 137.1 (36.9-329.1) | 1051581 (818771-1341385) | 33740 (15808-62365) | D |
|  | CXR-CAD | 211.2 (61-484.4) | 1055389 (824459-1344143) | 37548 (15183-70844) | D |
|  | Symptoms + CXR-CAD | 224.6 (66.1-512.5) | 1082400 (846666-1376055) | 64559 (36222-105301) | D |
|  | Pooled Xpert | 173.9 (49.5-405.2) | 1060174 (825542-1343333) | 42333 (20402-76436) | D |
| Exit | Symptoms | 19 (4.1-54) | 1028190 (800329-1312745) | 10349 (6163-16742) | ED |
|  | CXR-CAD | 31.2 (7.2-87.4) | 1032016 (804391-1316082) | 14175 (8199-23562) | ED |
|  | Symptoms + CXR-CAD | 33.6 (7.9-94.4) | 1038455 (808680-1324727) | 20614 (13120-31471) | ED |
|  | Pooled Xpert | 24.8 (5.7-68.9) | 1032982 (804583-1318178) | 15141 (9129-24301) | D |
| Annual | Symptoms | 275.1 (78.7-630.8) | 1053342 (816661-1340162) | 35500 (10089-72407) | D |
|  | CXR-CAD | 425.8 (132.8-954.1) | 1045096 (813419-1328612) | 27254 (-3512-72172) | Frontier |
|  | Symptoms + CXR-CAD | 452.9 (140.7-1016) | 1079097 (843251-1366883) | 61256 (27508-112081) | D |
|  | Pooled Xpert | 349.8 (106.1-789.4) | 1059282 (820309-1339507) | 41441 (10992-86095) | D |
| Biannual | Symptoms | 444.6 (132.1-1012.2) | 1080777 (843814-1372320) | 62936 (22277-117648) | D |
|  | CXR-CAD | 620.9 (199.8-1371.1) | 1060596 (835272-1341877) | 42755 (2133-93623) | Frontier |
|  | Symptoms + CXR-CAD | 648.6 (210.3-1433.8) | 1130588 (896735-1419920) | 112747 (62481-181052) | D |
|  | Pooled Xpert | 537.5 (167.6-1195.1) | 1086485 (851926-1366876) | 68643 (23427-128476) | D |
| Entry + exit + annual | Symptoms | 373.5 (109.4-866.6) | 1090072 (852274-1384766) | 72231 (33211-126147) | D |
|  | CXR-CAD | 535.9 (168.6-1192.4) | 1082419 (853899-1362927) | 64578 (22748-119507) | D |
|  | Symptoms + CXR-CAD | 563 (175.8-1241.3) | 1150246 (910081-1439005) | 132405 (77866-200403) | D |
|  | Pooled Xpert | 457.2 (139.1-1013.6) | 1103218 (863558-1391247) | 85377 (39630-150243) | D |
| Entry + exit + biannual | Symptoms | 511 (157.2-1168.4) | 1115344 (877428-1406670) | 97502 (44837-163716) | D |
|  | CXR-CAD | 679.1 (218.7-1494.1) | 1098123 (873835-1381391) | 80282 (29262-137914) | Frontier |
|  | Symptoms + CXR-CAD | 704.4 (230.5-1539.6) | 1203201 (966487-1496138) | 185359 (113691-272471) | Frontier |
|  | Pooled Xpert | 601.8 (188.1-1329.8) | 1127773 (893327-1411787) | 109932 (51529-182983) | D |

**Table S4. Optimal strategies by prison incidence.** Ranges of prison incidence under which each strategy has the highest probability of being the optimal strategy. N/A indicates that a given strategy was not optimal at any tested incidence level.

| **Optimal strategy (intervention, algorithm)** | **Incidence range (per 100,000 person-years)** | | |
| --- | --- | --- | --- |
|  | **Brazil** | **Colombia** | **Peru** |
| Base case | ≤ 116 | ≤ 121 | N/A |
| Annual screening, CXR-CAD | 117 - 214 | 122 - 297 | ≤ 443 |
| Biannual screening, CXR-CAD | 215 - 983 | 298 - 1080 | 444 - 1108 |
| Entry + exit + biannual, CXR-CAD | ≥ 984 | ≥ 1081 | 1109 - 5336 |
| Entry + exit + biannual, symptoms & CXR-CAD | N/A | N/A | ≥ 5337 |

**Table S5. Costs, effects, and cost-effectiveness of strategies on the efficiency frontier if CXR-CAD were unavailable.** Mean estimates are shown, standardized per 100,000 population. All estimates are population-wide (i.e., they include costs and effects accrued by the entire population, not just those in prison). Only strategies on the efficiency frontier are shown; all other strategies were dominated. Costs are in 2023 US dollars. DALYs, disability-adjusted life years; increm., incremental; ICER, incremental cost-effectiveness ratio.

| **Strategy (intervention, algorithm)** | **Cost (USD)** | **Effect (DALYs averted)** | **Increm. cost** | **Increm. effect** | **ICER** |
| --- | --- | --- | --- | --- | --- |
| Brazil | | | | | |
| Base case | 304676 | 0 | NA | NA | NA |
| Annual, symptoms | 340920 | 82 | 36244 | 82 | 440 |
| Biannual, symptoms | 378660 | 142 | 37739 | 59 | 634 |
| Biannual, pooled Xpert | 421364 | 175 | 42704 | 34 | 1270 |
| Entry + exit + biannual, pooled Xpert | 565428 | 215 | 144064 | 40 | 3607 |
| Colombia | | | | | |
| Base case | 233696 | 0 | NA | NA | NA |
| Annual, symptoms | 254494 | 81 | 20799 | 81 | 258 |
| Annual, pooled Xpert | 261038 | 101 | 6544 | 20 | 323 |
| Biannual, pooled Xpert | 290181 | 146 | 29143 | 45 | 649 |
| Entry + exit + biannual, pooled Xpert | 328027 | 161 | 37846 | 15 | 2551 |
| Peru | | | | | |
| Base case | 1017841 | 0 | NA | NA | NA |
| Annual, pooled Xpert | 1059282 | 350 | 41441 | 350 | 118 |
| Biannual, pooled Xpert | 1086485 | 538 | 27203 | 188 | 145 |
| Entry + exit + biannual, pooled Xpert | 1127773 | 602 | 41289 | 64 | 642 |

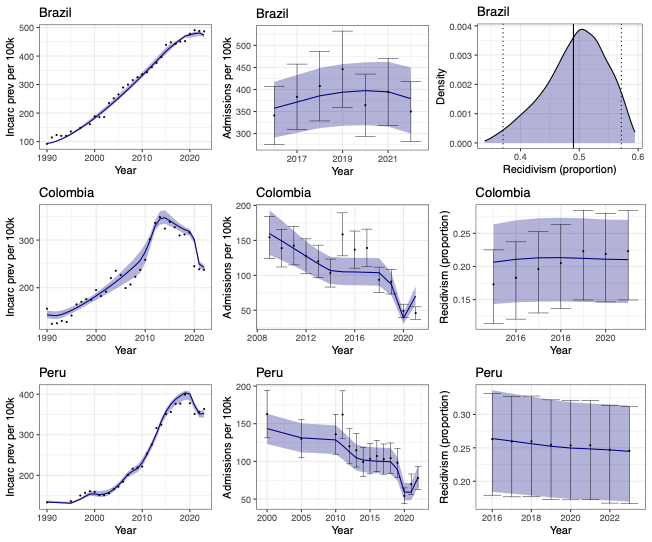

**Figure S1. Model fit to incarceration-related data targets.** Black points and error bars represent data

targets and 95% uncertainty bounds (if applicable), respectively. Dark blue lines and shaded bands represent median

model fits and 95% uncertainty intervals, respectively. In Brazil, recidivism data was only available for one year

(2013); the calibration target and uncertainty bounds are shown by the vertical black and dotted lines, respectively.

Incarc prev, incarceration prevalence; 100k, 100,000 population age 15+.

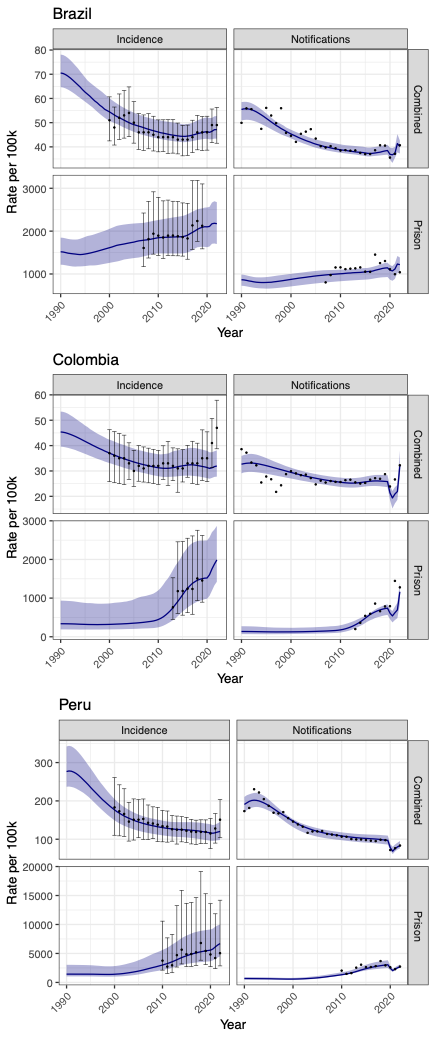

**Figure S2. Model fit to tuberculosis-related data targets.** Black points and error bars represent calibration

targets and 95% uncertainty bounds (if applicable), respectively. Dark blue lines and shaded bands represent median

model fits and 95% uncertainty intervals, respectively. “Combined” indicates population-wide notifications and incidence estimates. 100k, 100,000 person-years.

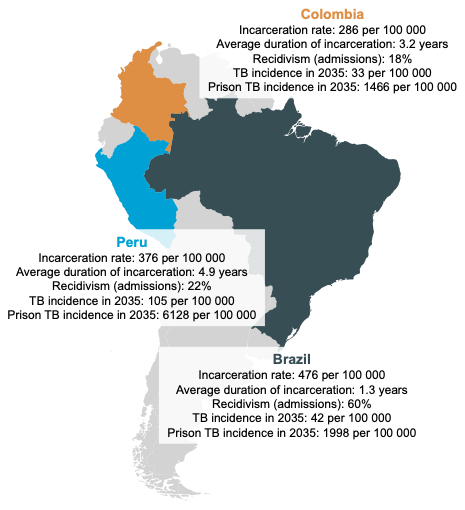

**Figure S3. Carceral characteristics and projected tuberculosis incidence in included countries.** Model projections for carceral characteristics at baseline and tuberculosis incidence in 2035 under the base-case scenario. Incarceration rates are for the population aged 15 and older. Recidivism refers to the proportion of people entering prison who have a prior incarceration history.

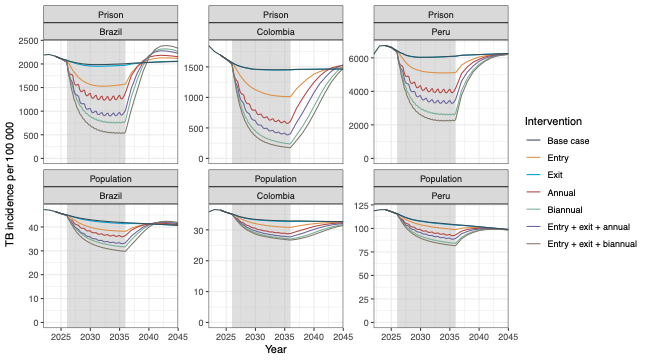
**Figure S4. Tuberculosis incidence over time under base-case and intervention scenarios**. Only interventions employing CXR-CAD are shown. Prison incidence is depicted in the top row; population-level incidence is in the bottom row. The shaded gray band shows the intervention period.

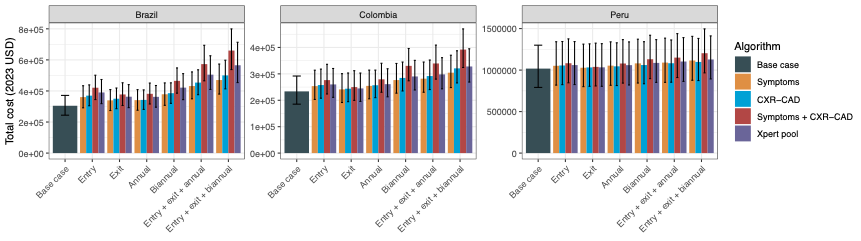

**Figure S5. Total costs of base-case scenario and screening interventions.** Costs are in 2023 US dollars (USD).

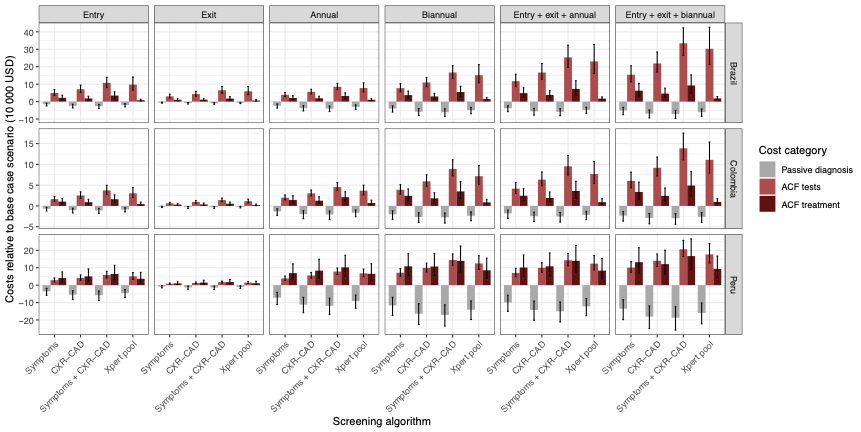

**Figure S6. Additional costs of screening interventions, relative to base-case scenario, disaggregated by costs of passive diagnosis, testing, and treatment.** Costs are in 2023 US dollars (USD). Costs of passive diagnosis include treatment costs for patients detected through passive diagnosis. ACF, active case finding.

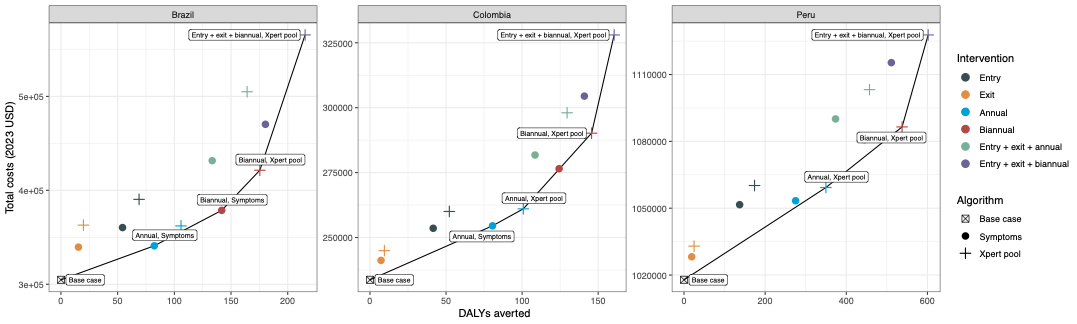

**Figure S7. Cost-effectiveness plane without algorithms using CXR-CAD.** Strategies on the efficient frontier are highlighted. Costs are in 2023 USD. DALYs, disability-adjusted life years.

6.	WHO. Global tuberculosis report 2024. 2024.

7.	Ministerio de Salud DGdM, Insumos y Drogas,. Estimación del umbral costo - efectividad para las evaluaciones económicas en salud. Informe técnico. Perú: 2022. (<https://repositorio-digemid.minsa.gob.pe/items/13bb3f97-69ab-49f7-8ec0-396c0d5416fb/full>).

8.	Ministério da Saúde. O uso de limiares de custo-efetividade nas decisões em saúde: recomendações da Comissão Nacional de Incorporação de Tecnologias no SUS. In: Secretaria de Ciência T, Inovação e Insumos Estratégicos em Saúde; Departamento de Gestão e Incorporação de Tecnologias em Saúde, ed. Brasilia/DF, Brazil2022.

9.	Espinosa O, Rodríguez-Lesmes P, Orozco L, et al. Estimating cost-effectiveness thresholds under a managed healthcare system: experiences from Colombia. Health Policy and Planning 2022;37(3):359-368. DOI: 10.1093/heapol/czab146.

10.	Ministerio da Saúde DATASUS. Casos de Tuberculose - Desde 2001 (SINAN). 2024.

11.	Santos AdS, de Oliveira RD, Lemos EF, et al. Yield, Efficiency, and Costs of Mass Screening Algorithms for Tuberculosis in Brazilian Prisons. Clinical Infectious Diseases 2021;72(5):771-777. DOI: 10.1093/cid/ciaa135.

12.	Pivetta de Araujo RC, Martinez L, da Silva Santos A, et al. Serial Mass Screening for Tuberculosis Among Incarcerated Persons in Brazil. Clin Infect Dis 2024;78(6):1669-1676. (In eng). DOI: 10.1093/cid/ciae055.

13.	Guerra J, Mogollón D, González D, et al. Active and latent tuberculosis among inmates in La Esperanza prison in Guaduas, Colombia. PLOS ONE 2019;14(1):e0209895. DOI: 10.1371/journal.pone.0209895.

14.	Ministerio de Salud (MINSA) Dirección de Prevención y Control de Tuberculosis (DPCTB). MINSA - DPCTB :: Portal de Información. 2024.

15.	IHME. Global Burden of Disease Study 2019. 2021.

16.	Menzies NA, Quaife M, Allwood BW, et al. Lifetime burden of disease due to incident tuberculosis: a global reappraisal including post-tuberculosis sequelae. The Lancet Global Health 2021;9(12):e1679-e1687. DOI: 10.1016/S2214-109X(21)00367-3.

17.	World Health Organization. Life tables by country. Global Health Observatory data repository2020.

18.	Pinto M, Steffen RE, Cobelens F, van den Hof S, Entringer A, Trajman A. Cost-effectiveness of the Xpert® MTB/RIF assay for tuberculosis diagnosis in Brazil. Int J Tuberc Lung Dis 2016;20(5):611-8. (In eng). DOI: 10.5588/ijtld.15.0455.

19.	Shah L, Rojas M, Mori O, et al. Cost-effectiveness of active case-finding of household contacts of pulmonary tuberculosis patients in a low HIV, tuberculosis-endemic urban area of Lima, Peru. Epidemiology and Infection 2017;145(6):1107-1117. DOI: 10.1017/S0950268816003186.

20.	Vesga JF, Mohamed MS, Shandal M, et al. The Return on Investment of Scaling Tuberculosis Screening and Preventive Treatment: A Modelling Study in Brazil, Georgia, Kenya, and South Africa. medRxiv 2024:2024.03.12.24303930. DOI: 10.1101/2024.03.12.24303930.

21.	Resch SC, Salomon JA, Murray M, Weinstein MC. Cost-Effectiveness of Treating Multidrug-Resistant Tuberculosis. PLOS Medicine 2006;3(7):e241. DOI: 10.1371/journal.pmed.0030241.
